# Low back pain in compensated Australian workers: a retrospective cohort study

**DOI:** 10.1101/2020.02.24.20027540

**Authors:** Michael Di Donato, Rachelle Buchbinder, Ross Iles, Shannon Gray, Alex Collie

## Abstract

**Objectives:** To describe incidence, duration, and patterns of working time loss claims in compensated Australian workers with low back pain (LBP), and compare this with other musculoskeletal conditions.

**Methods:** The National Dataset for Compensation-based Statistics (NDS) was used for this study. Any accepted workers’ compensation time loss claims for LBP, limb fracture or limb soft tissue disease occurring between July 2010 and June 2015 were included. Demographic information, occupation, and total cumulative time loss data were extracted. Counts, rates per 10,000 covered workers, the relative risk and median duration of time loss were calculated. Multivariate Cox and quantile regression models were used to determine factors affecting time loss duration and patterns.

**Results:** There were 56,102 LBP claims, 42,957 limb fracture claims, and 18,249 limb soft tissue disease claims. The relative risk of a claim for LBP was significantly greater than limb fractures after adjustment for all covariates (RR 1.30, 95%CI 1.29 – 1.32, p < 0.001). LBP claims had longer median time loss (9.39 weeks) than limb fracture claims (9.21 weeks). Quantile regression demonstrated that LBP claims were more likely than limb fracture claims to resolve within seven weeks, and to persist for periods beyond seven weeks.

**Conclusions:** There were differential patterns of time loss in LBP claims and limb fracture claims. The interaction between conditions, and policies and practices may contribute to these patterns. The findings should reiterate to workers’ compensation stakeholders the importance of returning a worker to work as soon as practicable, to avoid future delays or challenges.

**KEY MESSAGES:** *What is already known about this subject?:*

- LBP is a prevalent musculoskeletal symptom the leading contributor to the burden of disability for people of working age.
- Musculoskeletal conditions like LBP comprise the largest proportion of compensated conditions in Australia.

*What are the new findings?:*

- The incidence of claims for work-related LBP is higher than other work-related musculoskeletal conditions.
- Median time loss associated with a claim for LBP is similar to time loss for a claim for a limb fracture but less than for limb soft tissue disease claims.
- The likelihood of time loss duration is greater for workers with claims LBP than claims for limb fracture in the short term (seven weeks), but lower than limb fracture claims after this time.

*How might this impact on policy or clinical practice in the foreseeable future?:*

- Workers’ compensation stakeholders including clinicians, insurers, and workers’ compensation authorities should carefully consider the importance of returning a worker to work as soon as practicable to avoid future challenges, particularly for a condition as burdensome as LBP.

## INTRODUCTION

Low back pain (LBP) is a prevalent musculoskeletal symptom and the leading contributor to the burden of disease.^1-3^ Non-specific LBP typically has a favourable prognosis, with the majority of people with LBP recovering normal function within two weeks of pain onset.^3, 4^ However, LBP may be associated with activity limitation in the acute and even chronic setting.^3, 5-7^ A large proportion of LBP sufferers are of working age.^1-3^ A worker may seek wage replacement from an injury compensation or benefit system such as workers’ compensation or social security (hereafter referred to as ‘income support system’) if they are unable to work due to LBP.^8^

In Australia, if a condition such as LBP is determined to be work-related, then the worker may be eligible for payment for lost wages from a workers’ compensation scheme.^9, 10^ Injury and musculoskeletal disorders constitute the majority of workers’ compensation claims in Australia; 89% of serious claims in the 2016-17 financial year were for injuries and musculoskeletal disorders.^11, 12^ During the same period body stressing in the upper and lower back were the most common mechanism and location of injury, respectively.

While there have been previous estimates of the incidence of compensated musculoskeletal disorders in Australia,^12^ there are fewer estimates examining cases of work-related LBP specifically. Additionally, factors affecting compensated time loss have previously been estimated for musculoskeletal disorders more broadly, but not specifically for LBP in Australia.^13-15^ Finally, comparisons of claim incidence and time loss between claims for LBP and other musculoskeletal conditions have rarely been made in published literature.

The objectives of this study were therefore to (1) Determine the incidence of compensated work-related LBP in Australia and compare that with the incidence of two other common compensated work-related musculoskeletal disorders - limb fracture and limb soft tissue disease (muscle and tendon diseases); and (2) Determine and compare the duration of, patterns and factors affecting working time loss in compensated Australian workers with these three sets of conditions.

## METHODS

### Data source

This study used the National Dataset for Compensation-based Statistics (NDS). This administrative dataset is compiled by Safe Work Australia.^16^ Each state and territory in Australia has its own workers’ compensation scheme, and there are three Commonwealth (Federal) schemes.^10^ These systems are cause-based; wage replacement is only provided to those where an injury or illness is “attributable to a specific employment-related cause”.^9, 10, 17, 18^ Further description of the Australian workers’ compensation setting is available elsewhere.^9, 10^ These schemes provide data to Safe Work Australia to be collated in the NDS annually. The NDS contains claim-level data including: worker age, sex, occupation, industry, jurisdiction, socioeconomic status, remoteness, employment status, and cumulative time loss (reported in weeks). Safe Work Australia also supply a denominator dataset that contains the number of workers covered by workers’ compensation insurance in a given year for each of sex, age, occupation, and jurisdiction. The NDS and denominator data have been used in several previous studies.^19, 20^

### Sample

All accepted claims involving at least one day of compensated time away from work (i.e., time loss claims) by workers who sustained work-related (1) low back pain, (2) limb fracture, and (3) muscle and tendon diseases between 1 July 2010 and 30 June 2015 (i.e., five Australian financial years) were included. Each of the three groups were defined using the Type of Occurrence Classification System (TOOCS) version 3 revision 1 (see appendix 1).^21^The comparator groups were chosen as they represent musculoskeletal conditions that were hypothesised were likely to have different (limb fractures) and similar (muscle and tendon diseases) patterns of functional recovery and occupational outcomes compared to LBP.^22, 23^ The sample included workers aged > 15 years and < 80 years. Claims were excluded if they contained unlikely weekly working hours prior to a claim (< 1 hour and > 100 hours). Claims with time-loss less than two weeks or greater than 365 weeks were excluded, and censor indicator marked the maximum duration of any one claim at 104 weeks’ time loss (i.e., two years). ^19, 20, 24^ Filtering claims in this manner creates a standardised cohort across all jurisdictions. For example, some jurisdictions require an employer to pay the first 10 business days wage replacement (i.e., the two week filter), and each jurisdiction has a different maximum wage replacement period (i.e., censoring at 104 weeks, or two years). These eligibility criteria have been applied previously in studies using the NDS. ^19, 20, 24^ Application of these eligibility criteria is described in Figure 1.

**Figure 1.**
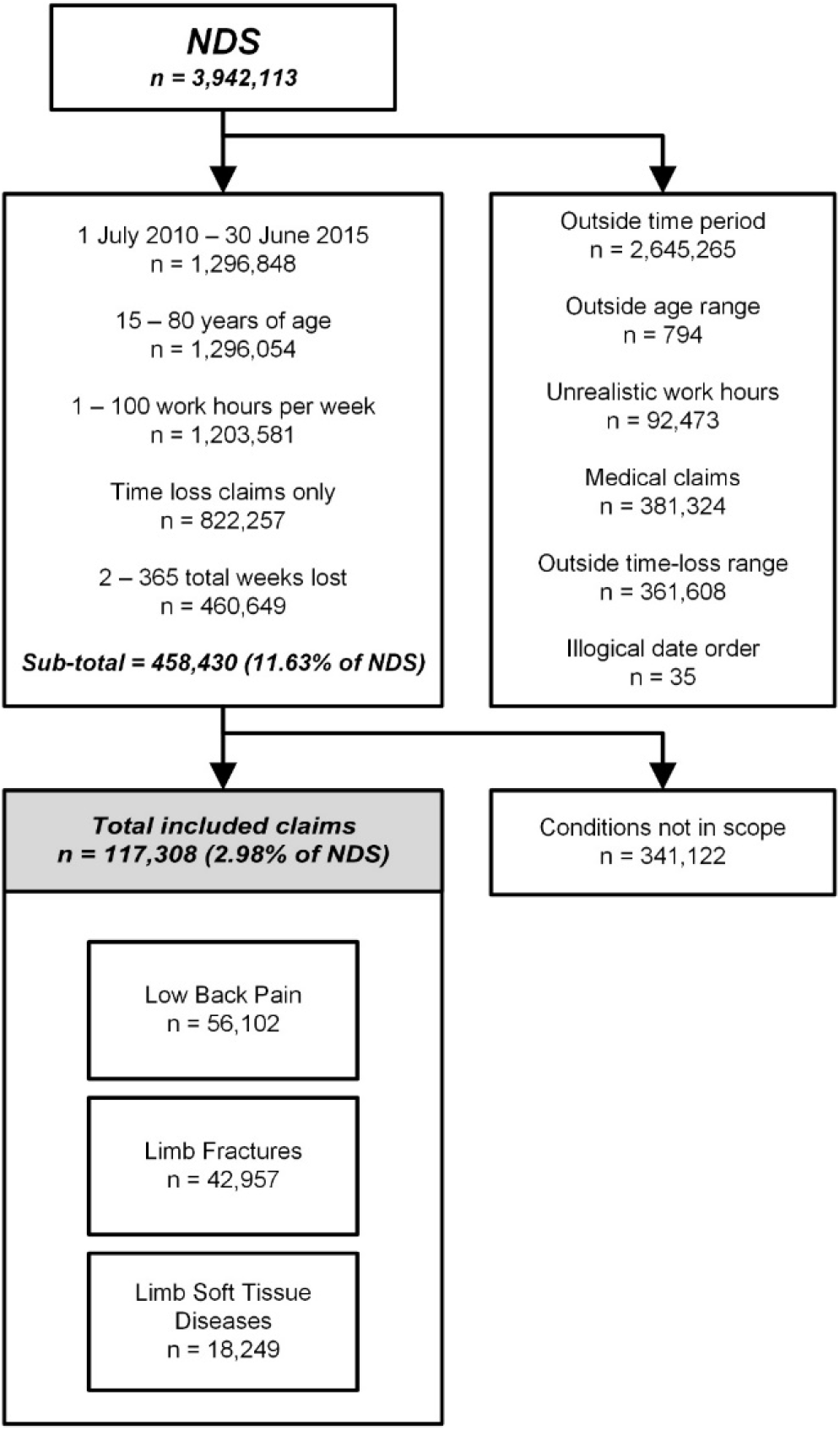
Sample selection process and number of included claims (NDS = National Dataset of Compensation-based Statistics)

### Outcome variables

The primary outcome variables for this study were (1) the incidence of accepted workers’ compensation time loss claims per 10,000 covered workers, and (2) duration of time loss for workers with accepted claims in weeks. These outcomes have been used in previous similar studies. 19, 20, 24

### Independent variable

Worker condition (i.e., LBP, limb fracture, or limb soft tissue disease) was the independent variable in analyses.

### Covariates

Covariates were chosen based on past association with study outcomes in groups of workers with LBP and other musculoskeletal conditions and availability within the dataset.^16, 19, 24^ Age at the time of injury was reported as age group (15-24 years, 25-34 years, 35-44 years, 45-55 years, and 55 > years), and sex reported in binary terms (male / female). Australian and New Zealand Classification of Occupations (ANZSCO) major codes were used to define eight major occupation groups.^25^ Jurisdiction was defined as the workers’ compensation jurisdiction in which the claim was accepted. Socioeconomic status was defined by the Index of Relative Socioeconomic Advantage and Disadvantage (IRSAD).^26, 27^ As per previous analyses, the middle three quintiles were collapsed, with the most advantaged and most disadvantaged quintiles at either extreme.^24^ Accessibility / Remoteness Index of Australia (ARIA) provided five categories of remoteness.^26, 28^ Finally, workers were classified as full-time if they worked ≥ 35 hours per week, or part-time otherwise.

Portions of the IRSAD (8.54%), ARIA (8.43%), and ANZSCO (0.39%) variables had missing values. IRSAD and ARIA are both derived from postcode, likely leading to similar proportion of missing values. Multivariate Imputation by Chained Equations (MICE) was used to impute the missing variables. MICE was performed for 10 imputations with 20 iterations of each imputation.^29^

### Analysis

Descriptive statistics were first used to determine the counts of time loss claims for LBP and the comparator conditions for each covariate. Claim incidence was calculated for LBP and the comparator conditions for sex, age group, occupation and jurisdiction. Poisson regression was used to calculate the unadjusted relative risk of a claim for each condition. The covariates sex, age group, occupation, and jurisdiction were then added to an adjusted model. Other covariates were not included in Poisson regression as denominator data (i.e., total covered workers) were not available. The log of the total number of covered workers was used as an offset in Poisson regression.^19^ Results were expressed as unadjusted and adjusted relative risk, with 95% confidence intervals (CI).

Descriptive statistics were used to determine the median and inter-quartile range (IQR) of time loss for LBP and the comparator conditions for all covariates. A Schoenfeld residual test indicated that the all conditions significantly violated the proportional hazards assumption.^30, 31^ A Cox proportional hazards model could therefore not be used to compare time loss between workers with LBP and the comparator conditions. However, to visualise the time loss differences between conditions a multivariate Cox model was created with the conditions as strata. This model was plotted as a survival curve, and demonstrated that workers with LBP and limb fractures appeared to have a fluctuating likelihood of cessation of time loss (i.e., a return to work) relative to one another (i.e., they were not proportional).

Quantile regression enabled a statistical comparison of time loss between LBP and the comparator conditions. A commonly used statistical method in econometrics, quantile regression performs regression within each of *tau* quantiles of the dependent variable, rather than across the entire dependent variable.^32, 33^ This method is ideal for analyses where the outcome variable is highly skewed and outliers may substantially affect the mean.^32, 33^ Both this analysis and other analyses of workers’ compensation data have consistently described the skewed nature of time loss as an outcome.^24, 34^ Quantile regression enables determination of how many weeks difference in time loss there are specific quantiles, and has also previously been used with this type of data^34^

Based on visualisation of time loss in the survival curves, the 10^th^, 25^th^, 50^th^, 75^th^, and 90^th^ quantiles were selected *a priori*. Analyses were performed with the entire imputed dataset, with all covariates included, in R 3.5 (Vienna, Austria),^35^ compiled in Microsoft Visual Studio 2017 (Redmond, USA).^36^

### Ethics

This study received ethics approval from the Monash University Human Research Ethics Committee (MUHREC) (Approval No. CF14/2995–2014001663, January 2019).

## RESULTS

### Incidence

There were a greater number of claims for LBP (n=56,102) than limb fracture (n=42,957) and limb soft tissue disease (n=18,249) (see Table 1 and Figure 1). The incidence of claims for LBP was also higher than both comparator conditions at 9.37 per 10,000 covered workers (7.17 per 10,000 covered workers for limb fracture, and 3.05 per 10,000 covered workers limb soft tissue disease). The relative risk of a claim for LBP was significantly greater than for limb fractures (RR 1.31, 95% CI 1.29 – 1.32, p < 0.001), and remained significant after adjustment for covariates (ARR 1.30, 95% CI 1.29 – 1.32, p < 0.001; ARR 0.43, 95% CI 0.42 – 0.44 for limb soft tissue disease).

**Table 1.**
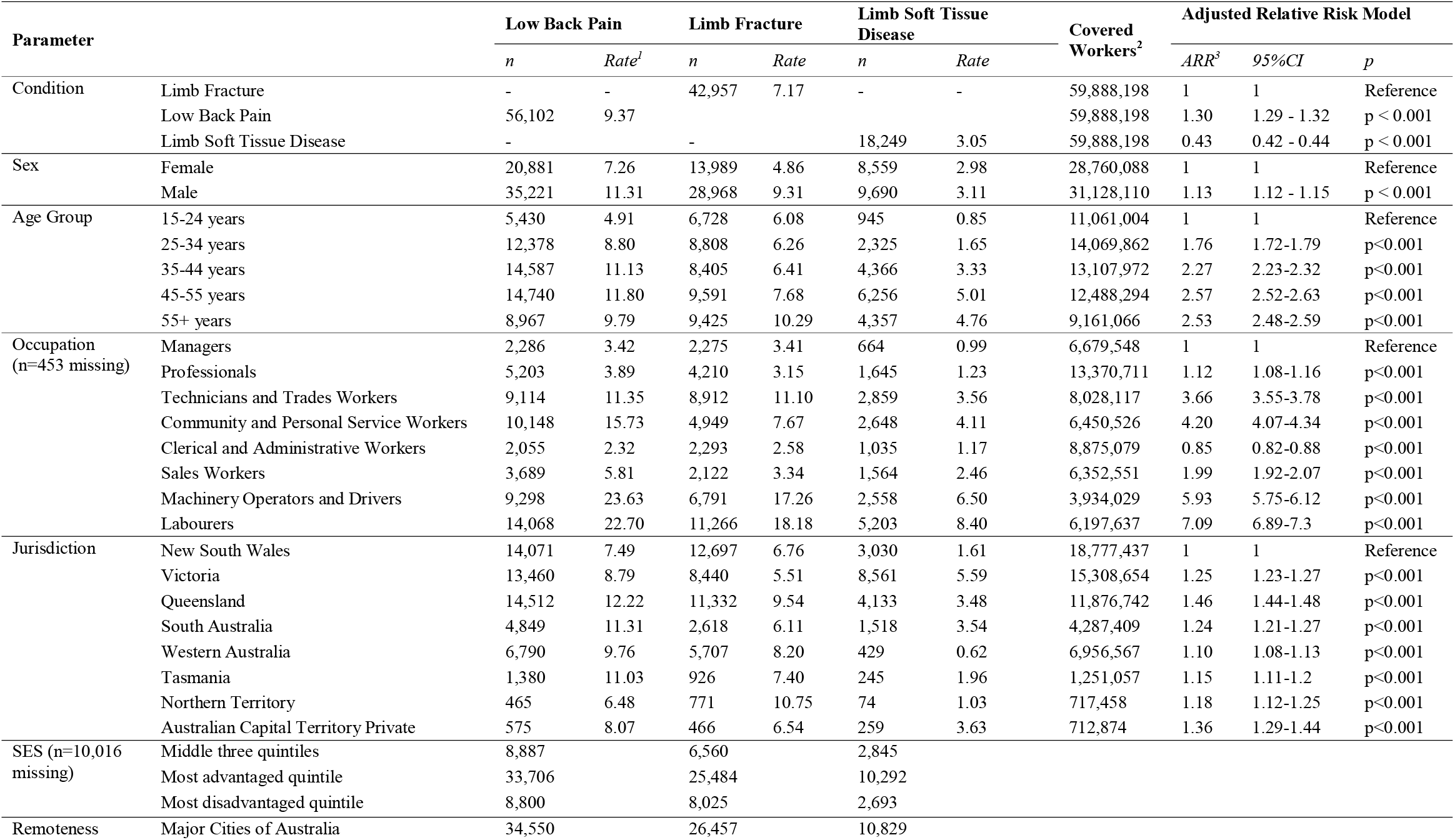
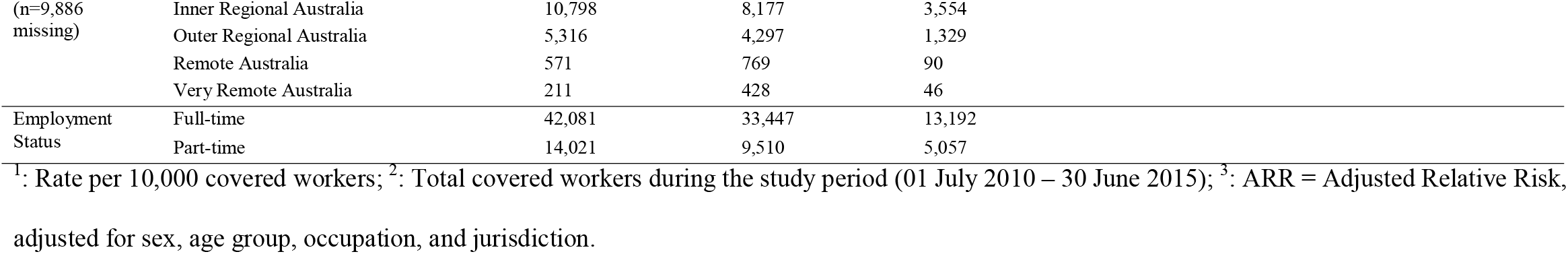
Incidence and relative risk of claims for included conditions between 01 July 2010 and 30 June 2015

The incidence of claims was greater for males than females for LBP and limb fracture claims, but there was a relatively small difference between sexes for limb soft tissue disease claims. The incidence of claims in all three condition groups generally increased with age, but the trend was most noticeable in LBP claims. More physically demanding occupations had a significantly greater incidence of claims for all conditions, and increased relative risk in the adjusted model. This was most notable for LBP claims, with *Machinery Operators and Drivers* and *Labourers* claiming for LBP at a rate of 23.6 and 22.7 per 10,000 covered workers, respectively.

The highest incidence of claims per jurisdiction was for LBP in Queensland (12.2 per 10,000 covered workers). However, other jurisdictions had relatively similar rates of LBP claims at 8.8 to 11.3 per 10,000 covered workers. The incidence of claims for limb fracture and limb soft tissue disease were lower, with limb soft tissue disease claims as low as 0.6 per 10,000 covered workers in Western Australia (WA).

### Time loss

Median time loss for LBP (9.39 weeks, IQR 3.95 – 30.2) and limb fractures (9.22 weeks, IQR 5.07 – 19.30) were similar, despite a more right-skewed 75^th^ percentile for LBP (see Table 2). The median time loss for limb soft tissue disease claims was greater at 14.40 weeks (IQR 5.92 – 40.4). A relatively large proportion of claims for LBP had a short duration of time loss (see Figure 2A). However, numerous claims lasted for a greater duration, extending the 75^th^ percentile. The majority of limb fracture claim time loss appeared to cluster closer to the median, with only a relatively small proportion of outliers. The distribution of time loss for limb soft tissue disease claims did not appear to follow a specific trend.

**Table 2.** Time loss and factors affecting time loss

| Variable |  | Low Back Pain |  | Limb Fracture |  | Limb Soft Tissue Disease |  |
| --- | --- | --- | --- | --- | --- | --- | --- |
|  |  | Median (weeks) | IQR | Median (weeks) | IQR | Median (weeks) | IQR |
| Condition | Limb Fracture | - | - | 9.21 | 5.07 - 19.30 | - | - |
|  | Low Back Pain | 9.39 | 3.95 - 30.20 | - | - | - | - |
|  | Limb Soft Tissue Disease | - | - | - | - | 14.40 | 5.92 - 40.40 |
| Sex | Female | 9.00 | 4.00 - 26.38 | 9.43 | 5.19 - 19.17 | 15.40 | 6.20 - 43.20 |
|  | Male | 9.60 | 3.80 - 32.8 | 9.20 | 5.03 - 19.4 | 13.80 | 5.60 - 38.60 |
| Age Group | 15-24 years | 7.00 | 3.39 - 19.14 | 7.30 | 4.40 - 13.20 | 8.80 | 4.00 - 23.81 |
|  | 25-34 years | 9.39 | 4.00 - 29.21 | 8.39 | 4.80 - 16.40 | 11.00 | 4.82 - 31.79 |
|  | 35-44 years | 10.00 | 4.00 - 34.00 | 9.59 | 5.00 - 20.59 | 13.79 | 5.71 - 40.60 |
|  | 45-55 years | 9.80 | 4.00 - 31.40 | 10.20 | 5.40 - 22.51 | 15.60 | 6.40 - 43.79 |
|  | 55+ years | 9.50 | 3.80 - 31.80 | 11.14 | 6.00 - 23.60 | 17.20 | 7.00 - 44.80 |
| Occupation | Managers | 10.60 | 4.39 - 37.55 | 8.82 | 4.94 - 18.47 | 14.70 | 6.77 - 45.58 |
|  | Professionals | 9.00 | 4.00 - 24.00 | 8.20 | 4.59 - 16.00 | 11.85 | 5.00 - 30.00 |
|  | Technicians and Trades Workers | 10.40 | 4.00 - 35.81 | 9.00 | 5.00 - 19.40 | 16.33 | 6.20 - 46.79 |
|  | Community and Personal Service Workers | 7.70 | 3.55 - 21.94 | 9.39 | 5.12 - 18.50 | 12.00 | 5.00 - 31.60 |
|  | Clerical and Administrative Workers | 8.75 | 3.60 - 25.95 | 8.80 | 4.61 - 17.00 | 12.40 | 5.45 - 29.40 |
|  | Sales Workers | 10.4 | 4.00 - 38.40 | 9.00 | 4.81 - 19.00 | 20.83 | 7.80 - 76.80 |
|  | Machinery Operators and Drivers | 10.31 | 4.00 - 35.80 | 10.2 | 5.60 - 22.00 | 15.20 | 6.19 - 42.21 |
|  | Labourers | 9.60 | 4.00 - 32.20 | 9.79 | 5.40 - 20.40 | 14.79 | 5.80 - 41.46 |
|  | Missing (n=453) | 9.40 | 4.00 - 17.80 | 7.00 | 4.20 - 15.20 | 8.83 | 5.60 - 24.92 |
| Jurisdiction | New South Wales | 6.77 | 3.20 - 21.00 | 8.60 | 4.60 - 17.60 | 9.80 | 4.04 - 25.00 |
|  | Victoria | 18.8 | 6.39 - 76.80 | 10.79 | 6.19 - 23.40 | 22.39 | 8.60 - 66.40 |
|  | Queensland | 7.80 | 3.60 - 19.00 | 8.88 | 5.00 - 18.00 | 10.25 | 4.53 - 23.00 |
|  | South Australia | 8.86 | 4.00 - 33.60 | 8.80 | 5.00 - 17.40 | 12.20 | 5.00 - 37.65 |
|  | Western Australia | 9.99 | 3.80 - 38.63 | 10.00 | 5.40 - 22.00 | 16.42 | 6.00 - 47.11 |
|  | Tasmania | 6.83 | 3.20 - 17.91 | 8.40 | 4.99 - 16.55 | 6.33 | 3.70 - 14.30 |
|  | Northern Territory | 11.33 | 4.20 - 34.60 | 9.80 | 4.80 - 21.27 | 16.90 | 5.85 - 32.55 |
|  | Australian Capital Territory Private | 6.80 | 3.44 - 18.21 | 9.86 | 5.20 - 21.40 | 9.68 | 4.19 - 23.92 |
| IRSAD | Middle three quintiles | 9.60 | 4.00 - 31.80 | 9.61 | 5.20 - 20.60 | 14.20 | 5.39 - 41.21 |
|  | Most advantaged quintile | 9.20 | 3.81 - 28.80 | 9.32 | 5.06 - 19.20 | 13.40 | 5.50 - 35.33 |
|  | Most disadvantaged quintile | 8.40 | 3.79 - 24.80 | 8.80 | 5.00 - 17.39 | 12.21 | 5.00 - 32.50 |
|  | Missing (n=10,016) | 13.00 | 4.60 - 55.35 | 10.00 | 5.23 - 23.00 | 30.40 | 10.24 - 100.00 |
| Remoteness | Major Cities of Australia | 9.32 | 3.95 - 29.10 | 9.20 | 5.00 - 19.20 | 13.60 | 5.60 - 36.39 |
|  | Inner Regional Australia | 9.00 | 3.80 - 28.49 | 9.20 | 5.12 - 18.80 | 12.80 | 5.00 - 34.00 |
|  | Outer Regional Australia | 8.39 | 3.76 - 26.00 | 9.30 | 5.17 - 19.60 | 13.00 | 5.00 - 36.20 |
|  | Remote Australia | 6.45 | 3.20 - 18.63 | 9.40 | 5.06 - 18.93 | 12.59 | 5.65 - 31.66 |
|  | Very Remote Australia | 8.00 | 3.49 - 27.22 | 9.47 | 4.80 - 17.40 | 11.00 | 3.89 - 30.03 |
|  | Missing (n=9,886) | 13.00 | 4.60 - 55.45 | 10.00 | 5.20 - 23.13 | 30.89 | 10.40 - 100.00 |
| Employment Status | Full-time | 9.58 | 4.00 - 30.40 | 9.20 | 5.00 - 19.00 | 13.60 | 5.79 - 36.00 |
|  | Part-time | 9.00 | 3.75 - 29.40 | 9.60 | 5.30 - 20.00 | 18.08 | 6.25 - 55.00 |

<sup>1</sup>: Adjusted for sex, age group, occupation, jurisdiction, socioeconomic status (SES), remoteness, and employment status – performed using imputed data

**Figure 2.**
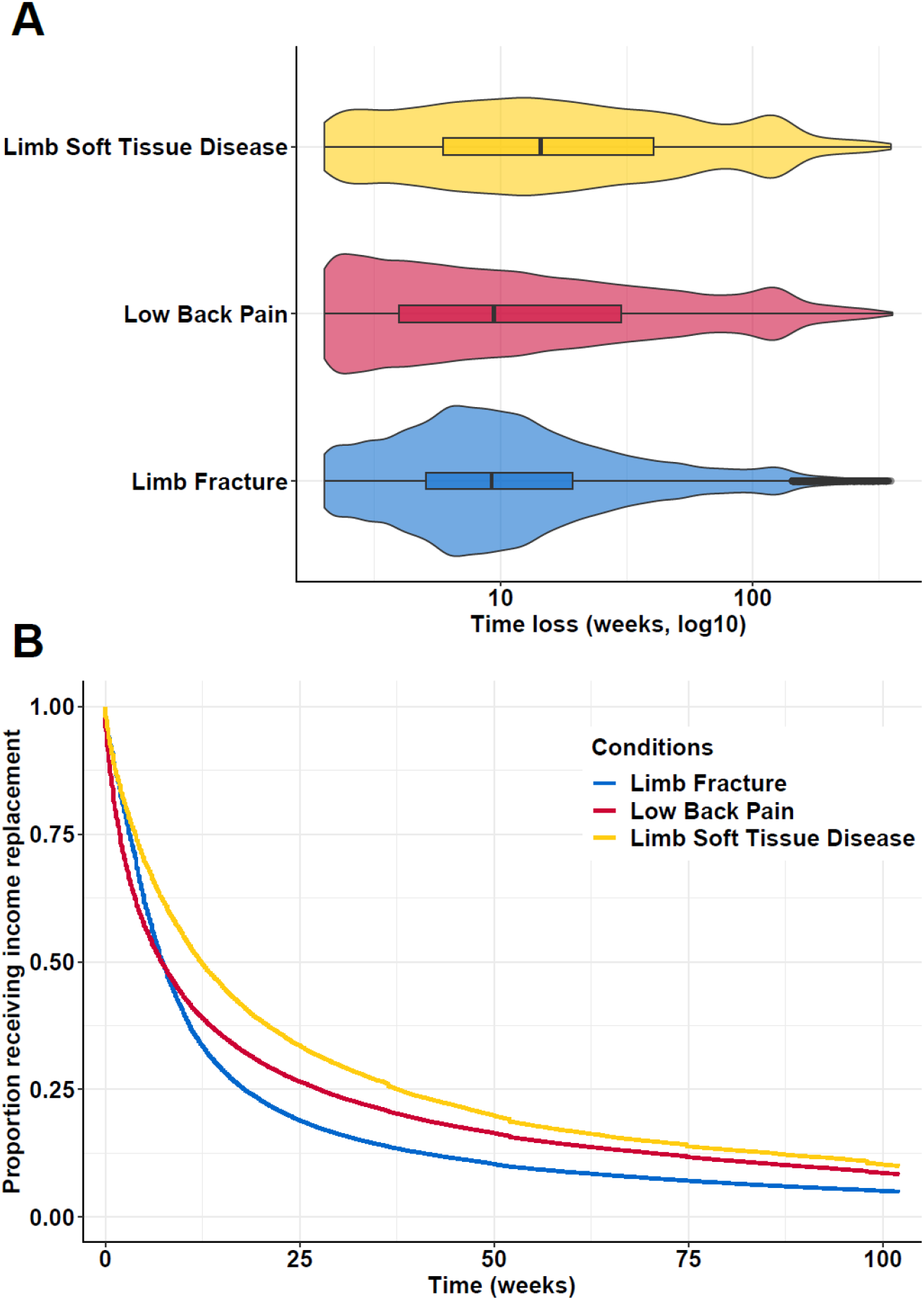
Visual representations of work-related time loss. (A) Violin and box plots demonstrating the distribution of claims by time loss, and median and IQR or time loss, by condition, and; (B) Cumulative survival curve of time loss (conditions are strata).

Median time loss was higher for males (9.60 weeks) than females (9.00 weeks) only for LBP. There did not appear to be an effect of age on time loss for LBP claims, nor were there substantial differences in time loss between occupations (see Table 2). Median time loss for LBP claims was highly varied between jurisdictions; LBP claims in NSW lasted a median of 6.77 weeks, compared to 18.80 weeks in Victoria. This same inter-jurisdictional variability was apparent for limb soft tissue diseases, but less so for limb fractures. There did not appear to be substantial differences in time loss for LBP claims between IRSAD. However, median time loss was up to 3.40 weeks greater for cases with missing socioeconomic status. LBP claims in major cities tended to have greater median time loss than in more remote areas.

Survival curves plotted based on the Cox model (see Figure 2B**Error! Reference source not found**.) demonstrated that a larger time loss for LBP claims were more likely to cease than limb fracture claims in approximately the six weeks. However, this trend reverses after approximately six weeks, with time loss for limb fracture claims more likely to cease after this time period. Limb soft tissue disease claims displayed the most shallow survival curve.

Quantile regression confirmed the patterns observed in the Cox model (see Table 3). LBP claims had approximately one week less time loss than limb fracture claims in the 25^th^ quantile of time loss (coef −1.12, 95%CI −1.20, −1.05). With all covariates set at reference values, claims for limb fracture at the 50^th^ quantile of time loss were modelled to last 7.46 weeks. There was no significant difference in time loss between LBP and limb fracture claims in the 50^th^ quantile (coef −0.14, 95%CI −0.31, 0.03). At the 90^th^ quantile time loss claims for LBP were 15.62 weeks greater than for limb fractures (95%CI 14.31, 16.94).

**Table 3.** Quantile regression analysis

| Quantile | Limb Fracture | Low Back Pain<sup>2</sup> |  |  | Limb Soft Tissue Disease |  |  |
| --- | --- | --- | --- | --- | --- | --- | --- |
|  | Weeks<sup>1</sup> | Coef (weeks)<sup>3</sup> | 95%CI | p | Coef (weeks) | 95%CI | p |
| 10th | 2.75 | -0.59 | (-0.63, -0.55) | 0.00 | -0.19 | (-0.28, -0.11) | 0.00 |
| 25th | 4.48 | -1.12 | (-1.2, -1.05) | 0.00 | 0.22 | (0.06, 0.38) | 0.01 |
| 50th | 7.46 | -0.14 | (-0.31, 0.03) | 0.12 | 3.18 | (2.8, 3.55) | 0.00 |
| 75th | 16.22 | 7.02 | (6.42, 7.61) | 0.00 | 9.63 | (8.6, 10.65) | 0.00 |
| 90th | 48.53 | 15.62 | (14.31, 16.94) | 0.00 | 12.29 | (10.87, 13.71) | 0.00 |

<sup>1</sup>: Calculated with all covariates set at reference values; <sup>2</sup>: Adjusted for sex, age group, occupation, jurisdiction, SES, remoteness, and employment status; <sup>3</sup>: coefficient and confidence interval are interpreted as additional / less weeks' time loss.

## DISCUSSION

This study aimed to determine the incidence and duration of working time loss due to work-related LBP in Australia compared with other common work-related musculoskeletal conditions. There was a greater incidence of LBP time loss claims compared to limb fracture and limb soft tissue disease claims. The relative risk of claiming for LBP was greater than claiming for limb fracture even after adjustment for demographic, occupational and injury characteristics. Time loss for LBP and limb fractures were relatively similar, with claims for limb soft tissue disease lasting longer. Distinct patterns of fluctuating likelihood of time loss emerged for LBP and limb fracture claims. Claims for LBP were significantly more likely to be resolved in the early phase (10^th^ and 25^th^ quantiles) than claims for limb fracture, but significantly less likely in the later phase (75^th^ and 90^th^ quantiles).

There were distinct patterns of time loss for each condition, despite similarities in median time loss. With the distribution of time loss clustered around the median (9.21 weeks), time loss for limb fracture claims appeared to follow what would be considered a typical clinical timeline of recovery for fracture healing.^23^ A large proportion of LBP claims appeared to have minimal time loss. This is promising, in that work-related LBP might follow expected recovery patterns for the majority of LBP cases – a typically positive prognosis and short-term disability.^3^ Visual patterns of time loss were confirmed statistically in the quantile regression model. LBP claims were more likely than limb fracture claims to resolve within seven weeks, and to persist for periods beyond seven weeks. The shift in likelihood of time loss duration may be a combination of factors from both conditions; workers with LBP may be more likely to return to work (i.e., time loss has ceased) as their pain has resolved quickly, with workers with limb fracture more likely to return to work after their fracture has healed sufficiently to resume activities. However, without further information it can only be hypothesised that this is the case.

The interaction between each condition and policies and practices may also contribute to the fluctuating time loss duration patterns observed in this analysis. For example, a practice such as diagnostic imaging may contribute to variable outcomes for each condition. Imaging has minimal diagnostic merit in the acute phase of LBP, and may be detrimental to a return to work for compensated workers.^37, 38^ In the case of limb fractures however, imaging is an essential and useful tool for diagnosis, and may be used to indicate that a worker is capable of returning to work. Other policy features may contribute to the observations made in this study. For example, both LBP and limb soft tissue disease claims observed a notable cluster of distribution of claims lasting approximately 100 weeks. This is unlikely to translate to recovery of large portion of claimants at this point. Instead, it is more likely due to the cessation of benefits in short-tail workers’ compensation schemes, such as Victoria.^10^

The higher rate of LBP in older workers aligns with previous research.^39^ However, the incidence of claims for work-related LBP does not necessarily reflect the incidence of work-related LBP. For example, lower claim rates in one jurisdiction do not necessarily mean that it is safer, and may instead indicate differences in system eligibility criteria, cultural or local norms, or health literacy.^24^ The higher rate of claims for LBP in more physically demanding occupations may indicate a link between physically demanding work and LBP. However, this link has previously been debated elsewhere.^40^ It may be that workers are more likely to lodge a claim for LBP if they have a physically demanding occupation because LBP limits their ability to perform their normal duties. Finally, the effect of policy differences on workers’ compensation claims rates between workers’ compensation schemes across Australia on time loss has previously been reported and is reflected in these findings.^24^

This study benefited from a large, population-wide sample. The use of robust and novel statistical methods provides new insights into time loss in this population. The analytic techniques used provide new prognostic insight in the return to work setting, by understanding the likelihood of time loss duration associated with each of the three included conditions at different times, rather than simply the pooled likelihood of return to work. There were also several limitations to this study. Firstly, there was missing data for some variables, and denominator data for all variables was not available.

The NDS is a secondary dataset that, despite extensive data cleaning and quality assurance, may contain errors. For example, it was possible that some conditions included in the data were misclassified. Injury classification (such as nature or location) is performed by workers’ compensation schemes, and may be subject to data entry errors. Several implausible conditions (e.g., tennis elbow in the hip) were identified and removed during data cleaning. Higher-level TOOCS codes (e.g., soft tissue diseases) were used to absorb this possible misclassification. However, this may have led to a cohort definition that was too clinically broad; an upper limb fracture and lower limb fracture are likely to have different effects on a person’s mobility. However, we believe that even with such limitations the analyses captured the overall important trends.

Finally, most statistical relationships were significant despite some relatively small effect sizes. This was likely due to the large sample size available for this study, and while not necessarily a limitation, future analyses may benefit from sub-sampling to ensure effects remain significant at even smaller sample sizes.

Future research should consider that time loss should not be viewed a single fixed value, but a fluctuating concept. Results presented here demonstrated that the likelihood of return to work may change with time depending on condition, and other recent research has demonstrated that gender has a similar time-dependent effect on working time loss outcomes.

As in the general population, LBP is a prevalent musculoskeletal symptom in compensated Australian workers. Time loss due to claims for LBP is relatively similar to claims for limb fractures. However, changes in the likelihood of time loss duration for LBP relative to limb fracture claims varies depending on the duration of the claim. These findings should reiterate to clinicians, claims managers, and workers’ compensation schemes the importance of returning a worker to work as soon as practicable, to avoid future delays and challenges.

## Data Availability

This study used the National Dataset of Compensation-based statistics (NDS). This dataset is compiled and administered by SafeWork Australia. The NDS is not available for distribution or access by the authors of this article.

## Acknowledgements

This paper uses data supplied by Safe Work Australia and has been compiled in collaboration with state, territory and Commonwealth workers’ compensation regulators. The views expressed are the responsibility of the author(s) and are not necessarily the views of Safe Work Australia or the state, territory and Commonwealth workers’ compensation regulators.

